# Protocol for developing the reporting guideline for the use of chatbots and other Generative Artificial intelligence tools in MEdical Research (GAMER)

**DOI:** 10.1101/2024.10.15.24315506

**Authors:** Xufei Luo, Yih Chung Tham, Mohammad Daher, Zhaoxiang Bian, Yaolong Chen, Janne Estill, the GAMER working group

**Author notes:** Correspondence to: Professor Yih Chung Tham, Department of Ophthalmology, Yong Loo Lin School of Medicine, National University of Singapore, Singapore., Professor Yaolong Chen, Evidence-Based Medicine Center, School of Basic Medical Sciences, Lanzhou University, No.199, Donggang West Road, Chengguan District, Lanzhou 730000, China., Professor Janne Estill, Evidence-Based Medicine Center, School of Basic Medical Sciences, Lanzhou University, No.199, Donggang West Road, Chengguan District, Lanzhou 730000, China.

## Abstract

**Background:** The integration of artificial intelligence (AI) has revolutionized medical research, offering innovative solutions for data collection, patient engagement, and information dissemination. Powerful generative AI (GAI) tools like ChatGPT and other similar chatbots have emerged, facilitating user interactions with virtual conversational agents. However, the increasing use of GAI tools in medical research presents challenges, including ethical concerns, data privacy issues, and the potential for generating false content. These issues necessitate standardization of reporting to ensure transparency and scientific rigor.

**Objective:** The development of the chatbots and other Generative AI tools in MEdical Research (GAMER) reporting guidelines aims to establish a comprehensive, standardized guideline for reporting the use of GAI tools in medical research.

**Methods:** The GAMER guidelines are being developed following the methodology recommended by the Enhancing the QUAlity and Transparency Of health Research (EQUATOR) network, involving scoping reviews and expert Delphi consensus. The process consists of three stages: preparatory work, Delphi survey, and testing and dissemination. The study is approved by the Ethics Committee of the Institute of Health Data Science at Lanzhou University (approval number: HDS-202406-01).

**Expected results:** The GAMER project was launched in July 2023 by the Evidence-Based Medicine Center of Lanzhou University and the WHO Collaborating Centre for Guideline Implementation and Knowledge Translation, and is scheduled to conclude in July 2024. The expected outcome of the GAMER project is a reporting checklist, accompanied by relevant terminology, examples, and explanations, to guide stakeholders in better reporting the use of GAI tools.

**Conclusion:** GAMER aims to guide researchers, reviewers, and editors in the transparent and scientific application of GAI tools in medical research. By providing a standardized reporting framework, GAMER seeks to enhance the clarity, completeness, and integrity of research involving GAI tools, promoting collaboration, comparability, and cumulative knowledge generation in AI-driven healthcare technologies.

**Trial Registration:** **We registered this protocol on the EQUATOR Network** (https://www.equator-network.org/library/reporting-guidelines-under-development/reporting-guidelines-under-development-for-other-study-designs/#CHEER).

## 1. Introduction

In recent years, the integration of artificial intelligence (AI) into research has revolutionized the landscape of medical research, offering innovative solutions for data collection, patient engagement, and information dissemination^1^. Powerful generative AI-based (GAI) tools like ChatGPT and other similar chatbots have emerged, facilitating interactions between users and AI through virtual conversational agents, and thereby presenting new opportunities for enhancing healthcare practice and research methodologgy^2^.

The increasing utilization of GAI tools linked to chatbots in medical research brings numerous opportunities and supports innovations, but also poses many challenges and issues. For example, the use of large language models may raise ethical concerns and data privacy issues^3-4^. Additionally, the generation of false content using large language models can constitute academic fraud^5^. These problems have detrimental effects on the academic community. Therefore, transparent and standardized reporting of the use of GAI tools is extremely important. Although many journals and academic institutions have some guidelines or suggestions on how to use GAI tools^6-10^, there is currently no universally recognized reporting guideline on how to transparently and scientifically report the use of GAI tools in medical research.

The chatbots and other Generative AI tools in MEdical Research (GAMER) aims to establish a comprehensive and standardized guideline for reporting essential elements in studies involving GAI based tools. The tool delineates key aspects such as whether GAI tools were used in the study and writing the paper, which tool and version were used, which sections of the paper it was applied to, the specific content areas it was used for, whether it was employed for language editing or content generation, and whether any manual testing was performed. By providing recommendations in which section each piece of information should be reported, GAMER seeks to create a reporting guideline that streamlines the reporting process, allowing researchers, practitioners, and policymakers to critically assess the quality and validity of studies involving chatbots interventions in medical research^3^.

The development of GAMER draws inspiration from established reporting guidelines in the field of medical research, such as the CONSORT (Consolidated Standards of Reporting Trials) statement for clinical trials^11^ and the STROBE (Strengthening the Reporting of Observational Studies in Epidemiology) statement for observational studies^12^. Adhering to GAMER will not only enhance the clarity and completeness of reporting in chatbot-related research but will also foster collaboration, comparability, and cumulative knowledge generation in the rapidly evolving field of AI-driven healthcare technologies. This article describes the methods and processes involved in the development of GAMER, providing a guarantee for the smooth completion and promotion of GAMER in the future.

## 2. Methods

We will follow the methodology recommended by the Enhancing the QUAlity and Transparency Of health Research (EQUATOR) website^13^ to develop the GAMER reporting guidelines, using methods such as scoping reviews and expert Delphi consensus to formulate the GAMER reporting checklist. The GAMER project commenced on July 1, 2023, and is scheduled to conclude by the end of July 2024. This study has been approved by the Ethics Committee of the Institute of Health Data Science, Lanzhou University (approval number: HDS-202406-01).

### 2.1 Project administration

The GAMER reporting guideline was initiated by the Evidence-Based Medicine Center of Lanzhou University, Chinese EQUATOR Center and the WHO Collaborating Centre for Guideline Implementation and Knowledge Translation. Dr. Yaolong Chen and Dr. Janne Estill are the co-project initiators, and Dr. Xufei Luo serves as the lead investigator and the main contact person for communication.

### 2.2 Funding sources

The project is funded by research project funds from the Research Unit of Evidence-Based Evaluation and Guidelines (2021RU017), Chinese Academy of Medical Sciences, School of Basic Medical Sciences, Lanzhou University, with no involvement of pharmaceutical companies or other third-party entities. The funding is used solely for relevant services, materials, travel, and publication expenses during the development process of GAMER.

### 2.3 Scope and target audience

The scope of GAMER is to assist stakeholders such as researchers, clinical doctors, editors, authors, peer reviewers, and other relevant parties in the smooth and barrier-free use and evaluation of GAI tools and similar chatbots for the purpose of composing medical research.

### 2.4 Development process

The development of GAMER reporting checklist consists of three stages (Figure 1). The first stage is the preparatory phase, involving the formation of an expert group, collection of initial items, and discussions on these items. The second stage primarily includes 1 to 2 rounds of Delphi survey and online face-to-face discussions, as well as multiple rounds of optimizing items. The third stage involves the publication of the checklist, as well as testing, dissemination and promotion of GAMER reporting guideline.

**Figure 1.**
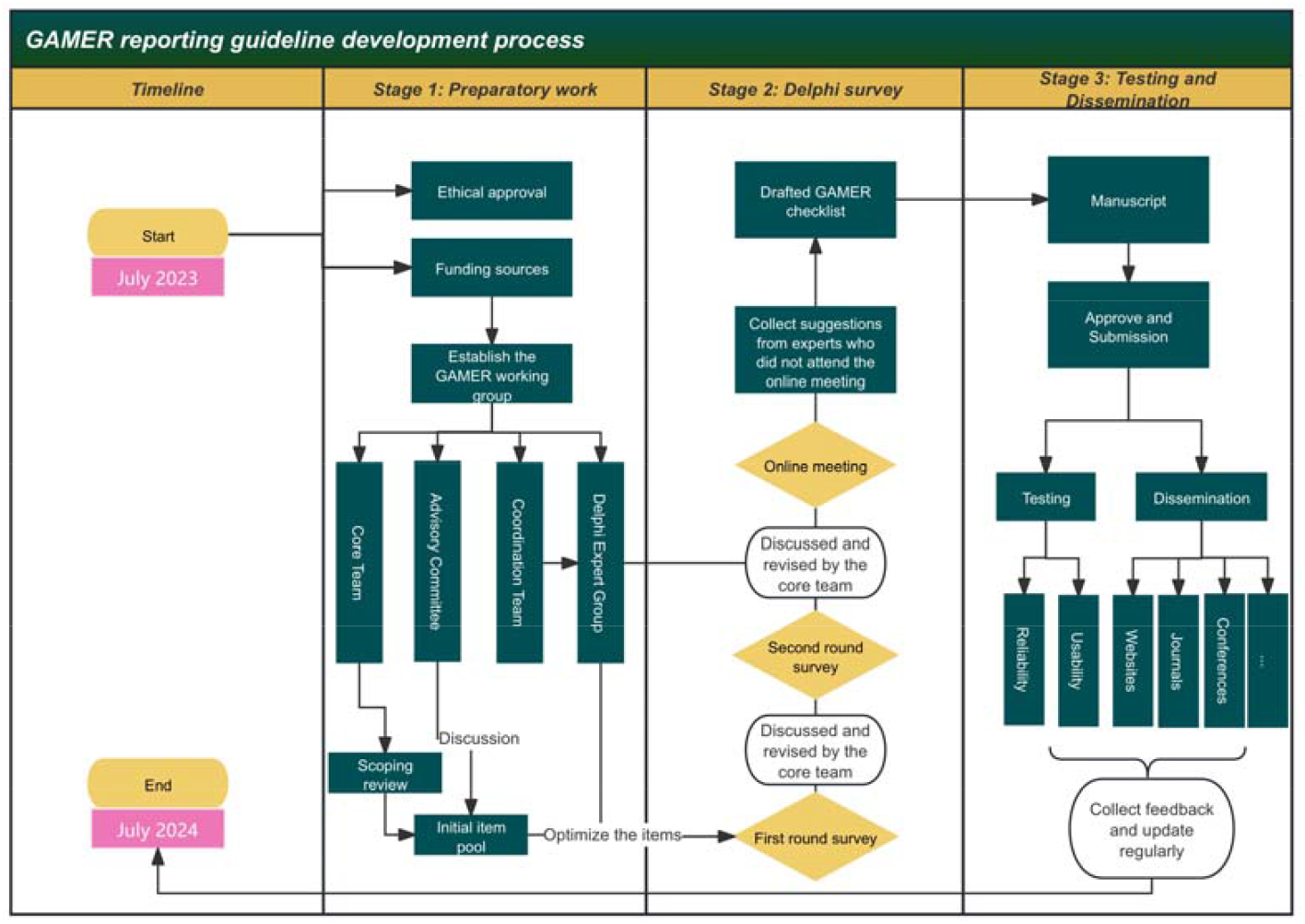
The three stages of development of the GAMER reporting guideline.

#### 2.4.1 Stage 1: Preparatory work

##### 1) Establishment of the Expert Panel

We plan to establish four expert groups. The first is the Advisory Committee, consisting of 3-5 authoritative experts familiar with GAI tools and experienced in the formulation of reporting guidelines. Their main responsibility is to provide advice on issues arising during the development of reporting guidelines. The second is the core team, primarily composed of members from the Evidence-Based Medicine Center at Lanzhou University. The group is responsible for conducting scoping reviews, generating the initial pool of items, revising the items, and drafting the final document. This group is leading team for GAMER development. The third is the Delphi expert group, comprised of experts with experience in a broad range of relevant disciplines from around the world. Inclusion criteria to be invited to this group are: 1) participation in the publication of GAI-related papers (obtained by searching PubMed for corresponding authors of relevant papers); 2) expertise in medicine-related disciplines; 3) willingness to participate in and support the development of GAMER; and 4) having no severe academic or financial conflicts of interests. The Delphi expert group’s responsibilities include proposing modifications to the GAMER checklist, voting, and determining the final modifications to the full document. The fourth one is coordination team including 3 to 5 members. This group is responsible for coordinating the development work, communicating with experts, organizing the meetings, sending emails, and organizing materials.

##### 2) Scoping review

We will perform a scoping review on GAI tools and similar chatbots, searching PubMed and Google Scholar to identify relevant articles. During the paper writing process, we will focus on literature related to the use of GAI tools like ChatGPT. We will examine the key elements reported in the published articles using GAI tools in medical research. Simultaneously, we reviewed guidelines for authors from top medical journals, as well as regulations from The International Committee of Medical Journal Editors (ICMJE) regarding the use of AI. This process aims to collect an initial pool of potential items for GAMER.

#### 2.4.2 Stage 2: Delphi survey

Once the preliminary items are completed, a survey will be disseminated through SurveyMonkey (http://surveymonkey.com) to reach consensus using a modified Delphi method. Anticipating one to two rounds but without a limit, the decision on the number of rounds depends on whether consensus is reached in the first round. Each initial item will be assigned a survey option ranging from 1 to 7 points, with 1 indicating strong disagreement and 7 indicating strong agreement. All statistics will be based on median scores. If the median score is between 1 and 3, the items will be excluded. If the score is 4 or 5 (or higher with substantial comments on the content), the item will be discussed and entered into the next round of the Delphi survey or the consensus meeting. If the score is 6 or 7 without any substantial comments, the item will be included in the final checklist. Online meetings will be held after the Delphi survey to collect the experts’ opinions and optimize and formulate the final checklist. All opinions and suggestions will be documented, and responses to each expert’s suggestions will be provided through email.

After the Delphi survey and virtual consensus meeting, we will refine and optimize the final items included. Once approved by the experts, we will prepare the manuscript for submission.

#### 2.4.3 Stage 3: Testing and Dissemination

After the GAMER checklist is finalized, we will invite authors of medical studies to use it when writing research papers involving GAI tools, simultaneously collecting their opinions and feedback to optimize and improve it. We will also invite experts from different medical disciplines to test the reliability and usability of GAMER checklist. Additionally, we will create explanatory documents and examples to assist users of GAMER in better understanding and implementing the checklist.

Furthermore, we plan to invite the consensus expert group members to translate GAMER into their local languages and promote the dissemination of the checklist. Additionally, we will actively reach out to journals, inviting them to feature GAMER on authors’ instructions.

Finally, we aim to establish a GAMER website to regularly update users on the latest developments and research findings related to GAMER reporting guidelines.

### 2.5 Timeline

The process and anticipated timeline for developing the GAMER reporting guideline are shown in Table 1.

**Table 1.**
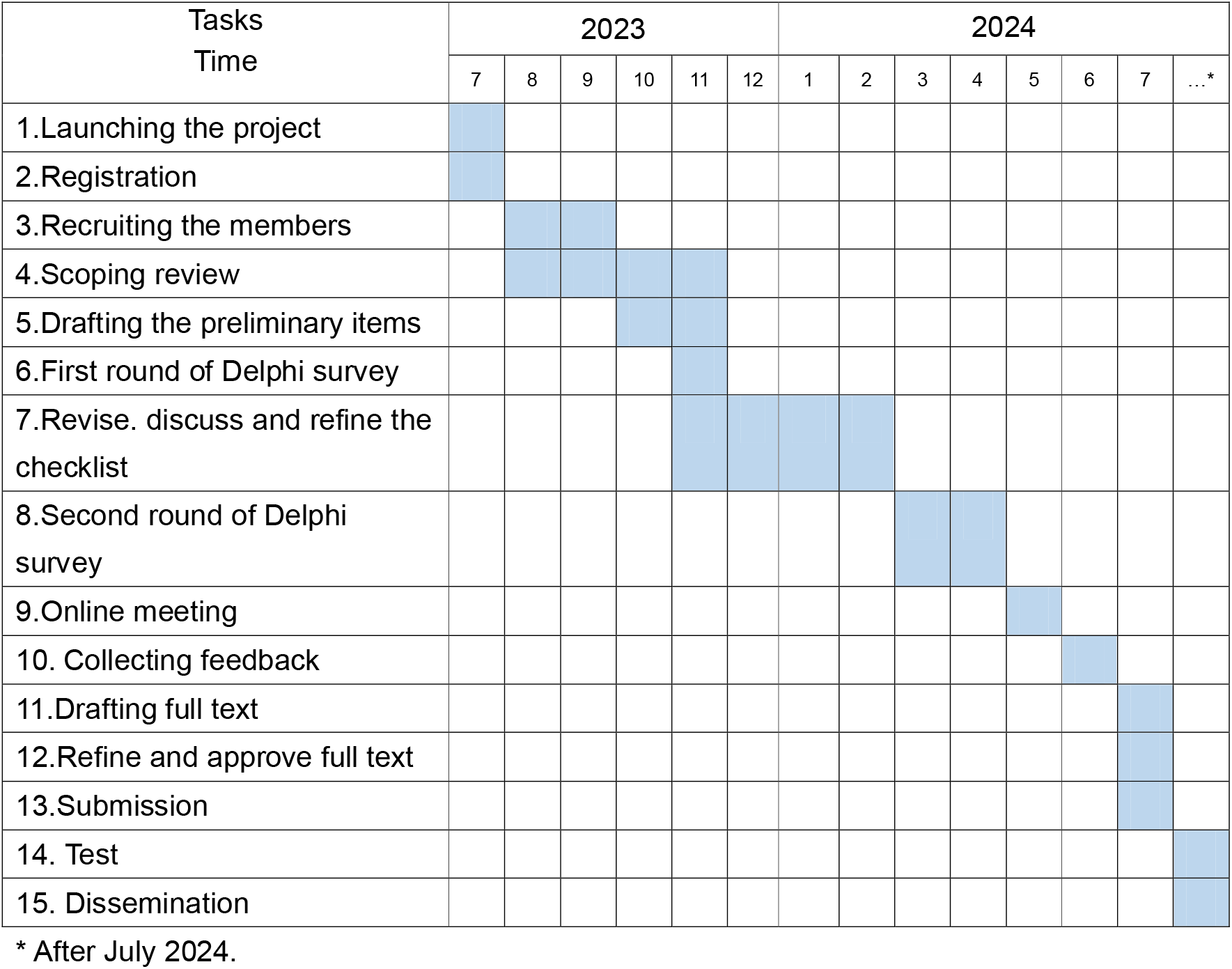
Timeline for GAMER development.

## 3. Discussion

We will follow the EQUATOR network recommended methodology to develop the GAMER reporting guidelines^13^, inviting experts from various disciplines and a broad geographical distribution to ensure the panel’s representativeness. We hope that GAMER can guide and assist those who use GAI tools in their research to report and apply these tools transparently and scientifically, thereby ensuring the integrity of the research. Additionally, we hope that GAMER will guide reviewers and editors in identifying and assessing manuscripts that have applied GAI tools, ensuring that the manuscripts meet the journal’s requirements for transparent reporting.

The development of GAMER is not only a response to the current utilization of GAI tools in medical research but also a forward-looking initiative. As AI technologies continue to evolve, the standardization introduced by GAMER will serve as a foundation for future advancements. The checklist’s scope, targeting various stakeholders including researchers, clinicians, editors, peer reviewers, and authors, positions GAMER as a versatile tool with far-reaching implications. The methodology, drawing from the EQUATOR guidelines, ensures a rigorous and evidence-based approach, further solidifying the credibility of GAMER.

The outlined plan for the dissemination and promotion of GAMER demonstrates a comprehensive strategy to ensure a widespread adoption of the reporting guidelines. By involving authors of medical research papers, collecting feedback, and actively engaging with journals, the GAMER project aims to integrate the checklist seamlessly into the publication process. The creation of a dedicated website further emphasizes the commitment to transparency and continuous improvement. This multifaceted approach is essential for the successful implementation of reporting guidelines, as it addresses both the supply side (researchers and authors) and the demand side (journals and users).

In conclusion, the GAMER project holds promise not only in addressing the immediate needs of standardizing the reporting of GAI tool use, but also in shaping the future landscape of medical research. The meticulous planning, expert involvement, and the strategic approach to dissemination position GAMER as a valuable resource for researchers, practitioners, and policymakers in the dynamic realm of AI-driven healthcare technologies.

## Data Availability

All data produced in the present study are available upon reasonable request to the authors.

## Authors’ Contributions

XL, ZB, YC, and JE designed this protocol. XL and YC drafted the manuscript. YCT, MD, YC, and JE critically reviewed the manuscript. All authors approved the final version for submission to the journal.

## Abbreviations

AI: Artificial Intelligence
ChatGPT: Chat Generative Pre-trained Transformer
GAMER: Chatbots and other Generative AI tools in MEdical Research
CONSORT: Consolidated Standards of Reporting Trials
EQUATOR: Enhancing the QUAlity and Transparency Of health Research
GAI: Generative Artificial Intelligence
ICMJE: The International Committee of Medical Journal Editors
STROBE: Strengthening the Reporting of Observational Studies in Epidemiology

## References

1. Chakraborty C, Pal S, Bhattacharya M, Dash S, Lee SS. Overview of Chatbots with special emphasis on artificial intelligence-enabled ChatGPT in medical science. Front Artif Intell. 2023 Oct 31;6:1237704. doi: 10.3389/frai.2023.1237704. PMCID: PMC10644239.

2. Wang X, Sanders HM, Liu Y, Seang K, Tran BX, Atanasov AG, Qiu Y, Tang S, Car J, Wang YX, Wong TY, Tham YC, Chung KC. ChatGPT: promise and challenges for deployment in low- and middle-income countries. Lancet Reg Health West Pac. 2023 Sep 15;41:100905. doi: 10.1016/j.lanwpc.2023.100905. PMID: 37731897; PMCID: PMC10507635.

3. Stahl BC, Eke D. The ethics of ChatGPT – Exploring the ethical issues of an emerging technology. International Journal of Information Management, 2024. 74, 102700. 10.1016/j.ijinfomgt.2023.102700

4. Wu X, Duan R, Ni J. Unveiling security, privacy, and ethical concerns of ChatGPT. Journal of Information and Intelligence, 2024, 2(2): 102–115.

5. De Angelis L, Baglivo F, Arzilli G, et al. ChatGPT and the rise of large language models: the new AI-driven infodemic threat in public health. Frontiers in public health, 2023, 11: 1166120.

6. Luo X, Estill J, Chen Y. The use of ChatGPT in medical research: do we need a reporting guideline? Int J Surg. 2023 Sep 14. doi: 10.1097/JS9.0000000000000737.

7. Flanagin A, Kendall-Taylor J, Bibbins-Domingo K. Guidance for Authors, Peer Reviewers, and Editors on Use of AI, Language Models, and Chatbots. JAMA. 2023;330(8):702-703. doi:10.1001/jama.2023.12500

8. Zielinski C, Winker MA, Aggarwal R, et al. Chatbots, Generative AI, and Scholarly Manuscripts. WAME Recommendations on Chatbots and Generative Artificial Intelligence in Relation to Scholarly Publications. WAME. May 31, 2023. https://wame.org/page3.php?id=106.

9. Alfonso F, Crea F. New recommendations of the International Committee of Medical Journal Editors: use of artificial intelligence. Eur Heart J. 2023;44(31):2888-2890. doi:10.1093/eurheartj/ehad448

10. Castellanos-Gomez A. Good Practices for Scientific Article Writing with ChatGPT and Other Artificial Intelligence Language Models. Nanomanufacturing. 2023; 3(2):135-138. doi:10.3390/nanomanufacturing3020009.

11. Moher D, Hopewell S, Schulz KF, Montori V, Gøtzsche PC, Devereaux PJ, Elbourne D, Egger M, Altman DG. CONSORT 2010 explanation and elaboration: updated guidelines for reporting parallel group randomised trials. BMJ. 2010 Mar 23;340:c869. doi: 10.1136/bmj.c869.

12. von Elm E, Altman DG, Egger M, Pocock SJ, Gøtzsche PC, Vandenbroucke JP; STROBE Initiative. The Strengthening the Reporting of Observational Studies in Epidemiology (STROBE) statement: guidelines for reporting observational studies. Ann Intern Med. 2007 Oct 16;147(8):573–7. doi: 10.7326/0003-4819-147-8-200710160-00010.

13. Moher D, Schulz KF, Simera I, Altman DG. Guidance for developers of health research reporting guidelines. PLoS Med. 2010 Feb 16;7(2):e1000217. doi: 10.1371/journal.pmed.1000217. PMID: 20169112; PMCID: PMC2821895.

